# HORMONE EXPOSURE AND VENOUS THROMBOEMBOLISM IN COMMERCIALLY-INSURED WOMEN 50 TO 64 YEARS OF AGE

**DOI:** 10.1101/2022.11.19.22282547

**Authors:** Susan C. Weller, John W. Davis, Laura Porterfield, Lu Chen, Gregg Wilkinson

**Author notes:** Corresponding Author: Susan C. Weller, Dept Population Health Science, School of Public and Population Health, University of Texas Medical Branch, 300 Harborside Dr., Galveston, Tx 77555-1153. **Competing interests** None declared. **Funding** The data were obtained with a grant to SCW from the Texas Academy of Family Physicians Foundation.

## Abstract

**Objective:** Determine whether hormone-associated venous thromboembolism (VTE) risk varies by exposure route and formulation in 50-64 year-old US women.

**Design:** Nested case-control study.

**Setting:** Large US commercially-insured population with patient-level claims data.

**Participants:** Women aged 50-64 years with at least one year of enrollment. Controls were matched to incident cases (10:1) on VTE date and case’s age (+/− 2yrs). Exclusions included prior VTE, intravascular vena cava (IVC) filter within twelve months, and anticoagulant exposure within 14 days.

**Exposures:** All estrogen and progestogen prescriptions (with route and formulation) filled within 12 months prior to index date were coded as current (0-60 days), past (61-365 days), or none. Contraceptives were categorized separately.

**Outcome:** Acute VTE cases were identified with ICD codes plus anticoagulant, IVC filter, or death within 30 days.

**Results:** Conditional logic regression analyses controlled for differences between cases (n=20,359) and controls (n=203,590) in Elixhauser comorbidities and VTE risk factors. Odds ratios (OR) were as follows: for current oral, unopposed estradiol 1.24 (95% CI: 1.09 to 1.40) or conjugated equine estrogen (CEE) 1.46 (95% CI: 1.28 to 1.68); for progestogens with estradiol 1.14 (95% CI: 0.95 to 1.37), with CEE 1.52 (95% CI: 1.25 to 1.84), or with ethinyl estradiol 2.35 (95% CI: 1.71 to 3.25). Current transdermal estradiol had the lowest ORs, whether unopposed, 0.70 (95% CI: 0.59 to 0.83) or combined with progestogens, 0.73 (95% CI: 0.56 to 0.96), but varied by progestogen. The OR for estrogen-progestogen contraceptives was 5.22 (95% CI: 4.67 to 5.84) compared to no exposure and 4.24 (95% CI: 3.64 to 4.98) compared to combined MHT.

**Conclusions:** In 50-64-year-old women, transdermal menopausal hormone therapy (estradiol with or without progestogens) did not elevate VTE risk. In contrast, contraceptives markedly increased VTE risk.

**Summary Boxes:** 

**What is already known on this topic?:**

- Randomized controlled trials indicate that relative risk for venous thromboembolism (VTE) is approximately twice as high with menopausal hormone therapy (MHT) containing conjugated equine estrogen (CEE) with or without medroxyprogesterone acetate compared to no hormone exposure.
- Recent large, observational studies in the UK and Europe suggest that estradiol is lower risk than CEE and transdermal estradiol does not raise VTE risk compared to no hormone exposure, but results may not generalize to the United States because of differences in formulary, prescribing patterns, and background VTE incidence.

**What this study adds:**

- Using a large medical record database for US commercially-insured women 50-64 years of age, results confirmed that VTE risk was higher for oral compared to transdermal MHT and transdermal MHT (unopposed estrogen or combined with a progestogen) did not increase risk for VTE compared to no hormone exposure. However, unique US prescribing patterns included MHT with transdermal estradiol plus oral progestogens and MHT with ethinyl estradiol.
- MHT estrogen formulation affected VTE risk: ethinyl estradiol had higher risk than CEE, and CEE had higher risk than estradiol.
- Combined hormonal contraceptives (oral, vaginal, transdermal) had a markedly higher increase in VTE compared to MHT.

---

Two decades ago, the Women’ s Health Initiative (WHI)[1,2] and the Heart and Estrogen-Progestin Replacement Study (HERS)[3,4] found that menopausal hormone therapy (MHT) lacked cardiovascular benefit and increased incidence of thrombotic events, particularly venous thromboembolism (VTE). Oral conjugated equine estrogen (CEE) approximately doubled the relative risk of VTE, with slightly lower risk when initiated closer to menopause.[5],[6] New evidence on VTE risk from UK and European observational studies suggests that hormone type (estrogen alone or combined), route, and formulation may affect risk for adverse outcomes.[7–13] Specifically, results suggest that VTE risk is lower for estradiol than for CEE and that transdermal MHT does not raise risk.

The question is whether these results replicate in other national contexts. US formularies, prescribing patterns, and VTE incidence differ from those in Europe. For example, dydrogesterone, a lower risk progestin, is unavailable and US incidence of VTE is much higher than in Europe.[18] The WHI and HERS results vastly reduced use of MHT,[14–16] but approximately eight percent of US women 50 and older continue to use non-contraceptive estrogens,[16] and little data exist as to how many additional women in this age range use hormonal contraceptives. Common indications for MHT include hot flashes, vaginal dryness, disruptions in sleep patterns, and inability to concentrate. However, many women who suffer from menopausal symptoms are not prescribed MHT because US clinical guidelines discourage its use.[17]

This study examined VTE risk from hormone exposures within the US context. Using a large, US population-based administrative health database, risk was assessed in commercially-insured US women 50-64 years of age without a prior VTE. A nested case-control design with matched cases and controls estimated VTE risk for hormone exposures by administration route and formulation, while controlling for other risk factors. The study focused on women entering the menopausal period, as they may be most likely to desire and use MHT.

## METHODS

### Data

The OPTUM de-identified Clinformatics^®^ Data Mart Database contains patient-level, longitudinal billing data for one of the largest US commercially-insured populations. This de-identified longitudinal database of administrative, medical, pharmacy, and laboratory claims represents approximately 62M unique members (2007-2018) across all US regions.

A base cohort of women 50-64 years of age was selected based on the median age of menopause at 49.6 years[19] and the large transition of enrollees from commercial insurance to Medicare at age 65. Women with at least one year of enrollment between January 1, 2007 to December 31, 2019 were eligible.

#### Patient and Public Involvement

The limited funding for this project focused on obtaining a commercial dataset of anonymised secondary data. Patients were not identified or contacted, nor were they involved in developing the research question, selection of the outcome measure, or study design. Results have been disseminated through continuing medical education efforts to providers, e.g. the Texas Association of Family Practice.

### Incident Cases

Using the base cohort, acute VTE diagnoses were identified with ICD codes (DVT: ICD9: 451.1x, 451.2x, 451.81, 451.9x, 453.4x; ICD10: I80.1x, I80.2x, I80.3, I80.9, I82.4x, I82.90; PE: ICD9: 415.1x; ICD10: I26.0x, I26.9x), regardless of the setting of the diagnosis (inpatient or outpatient). Cases required a VTE diagnosis plus filling of an anticoagulant prescription (excluding heparin flushes), placement of an intravascular vena cava (IVC) filter, or death within 30 days following diagnosis to minimize inclusion of suspected or unconfirmed cases. VTE case identification in administrative claims data with this method has shown good accuracy.[20] Cases had to occur after 12-months continuous enrollment without a VTE or inferior vena cava (IVC) filter; date of VTE defined the index date. Those with anticoagulant long-term use (in past 12 months) or exposure within 14 days of the index date (heparin flushes excepted) were excluded.

### Controls

Controls were matched to incident cases (10:1) on index date and age (+2 years) and selected at random. A minimal matching strategy was used to increase generalization to the population. Controls had to be continuously enrolled in the 12 months prior to the index date with the same exclusion criteria as cases.

### Hormone Exposures

For cases and controls, all prescriptions filled within the year prior to the index date containing estrogen and/or progestogen were recorded, as well as hormone type (unopposed estrogen, estrogen-progestogen combinations, progesterone only), administration route (oral, transdermal, vaginal), estrogen formulation (estradiol, CEE, ethinyl estradiol, etc.), and progestogen formulation (progesterone, norethindrone, medroxyprogesterone acetate/MPA, etc.). Low-dose vaginal estrogens used exclusively for treatment of local genitourinary symptoms were excluded due to their lack of effect on systemic estradiol levels.[21] Exposure was coded as the most recent exposure: within 30 days, 31-60 days, 61-90 days, or 91-365 days prior to the index date. All 30- and 90-day prescriptions were considered for subsequent 30- or 90-day exposure periods. Estrogen-progestogen contraceptives were categorized separately. Those without a hormonal prescription in the previous year were considered unexposed and formed the reference category.

Women with overlapping prescriptions for an estrogen and a progestogen (e.g., transdermal estrogen with an oral progestogen) were considered exposed to both. For women exposed to estrogen through two different routes of administration (oral and either transdermal or vaginal) in the same month, the exposure route was considered oral.

### Covariates

Known VTE risk factors were recorded: hospitalization/surgery or trauma in the previous 30 days; malignancy, varicose veins, hypercoagulable conditions, and smoking documented in previous 12 months. In addition, coronary artery disease (CAD), stroke, and Elixhauser comorbidities (31 conditions such as hypertension, diabetes, and liver disease that are predictive of mortality) [22,23] in the past 12 months were included to control for general health status between cases and controls (possible sources of indication bias).

### Analysis

Relative risks for hormone exposures were estimated with odds ratios (ORs) and 95% Wald CIs from conditional logistic regression models. Analyses were adjusted for known VTE risk factors, CAD, stroke, Elixhauser comorbidities, age, and US region of residence. The Elixhauser comorbidities were reduced into a single index: none, one or two, and three or more. Cancers and coagulopathy were removed from the Elixhauser index and analyzed as separate variables as known VTE risk factors. Models explored timing of hormone exposure to identify the highest risk exposure period (0-30, 0-60, or 0-90 days prior to the index date). Separate models assessed risk associated with type of therapy (estrogen with/without other hormones), route of administration, and formulation while controlling for other risk factors. Stratified analyses explored possible subsample differences, first by excluding anyone with cancer or hypercoagulable conditions and second by stratifying on age (<58, ≥58) as most women reach menopause by 58. All statistical analyses were performed with SAS Version 9.4.[24] Number needed to harm (NNH) [25] was estimated assuming a population VTE incidence of 180/100,000 for women in this age range[18] and the adjusted odds ratios from conditional logistic regression models.

## RESULTS

### Sample

From the base cohort of women, 20,359 incident cases of acute VTE met inclusion criteria and were matched with 203,590 controls (Figure 1). Among cases, 76% were primary diagnoses, 89.37% (18,194) received anticoagulant therapy, 12.00% (2,444) had an IVC filter placed, and 5.88% (1197) died within 30 days of diagnosis. There were 10,995 (54.01%) cases of PE (with or without DVT) and 9,364 (45.99%) cases of DVT (without PE). Cases had more comorbidities and risk factors than controls: 55.94% of cases and 22.45% of controls had three or more Elixhauser comorbidities; 26.00% of cases and 5.59% of controls had cancer in the previous year, and 30.22% of cases and 2.29% of controls were recently hospitalized or had surgery (Table 1).

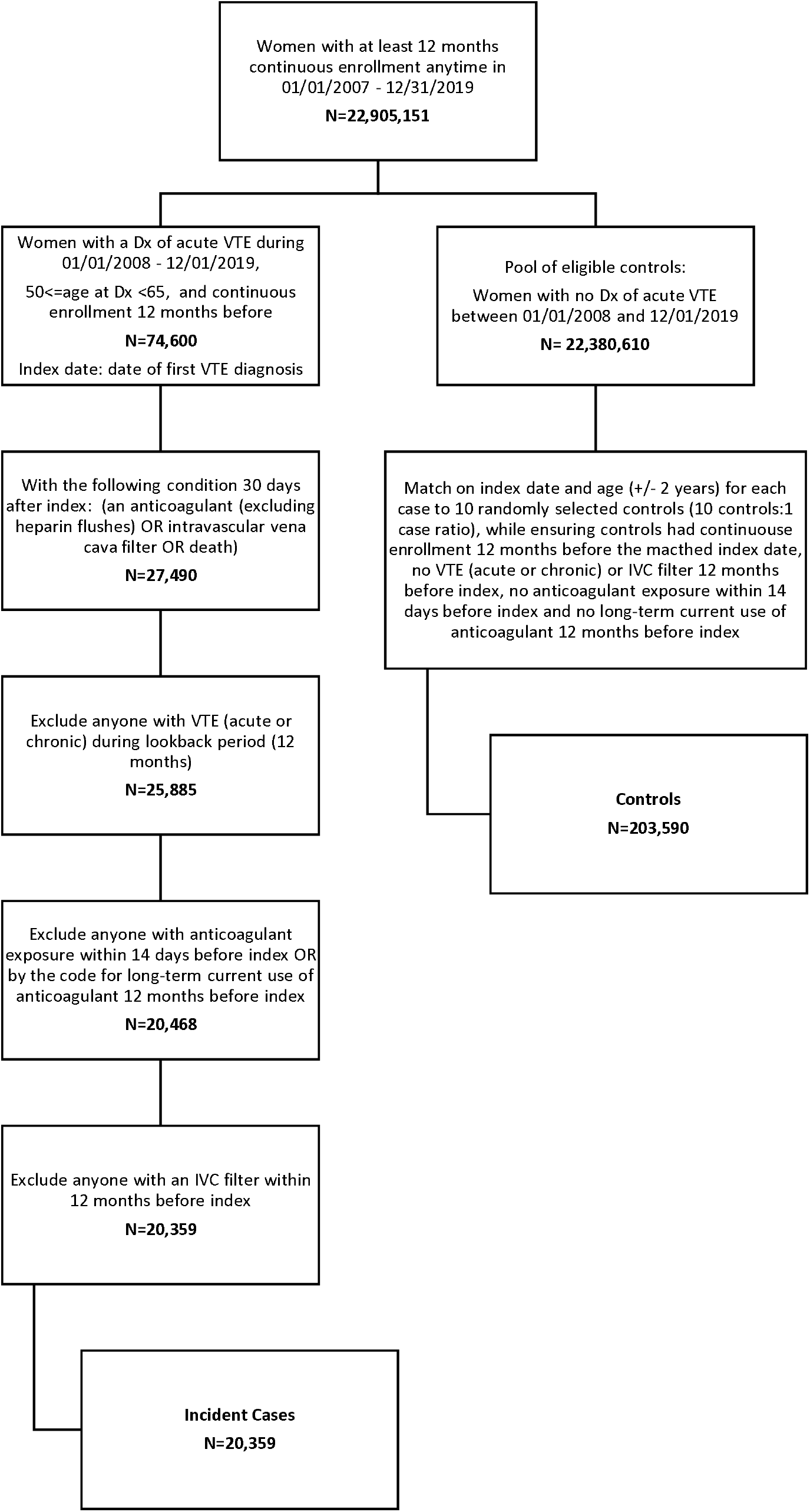

**TABLE 1:** COVARIATES.

| **Variable** | **VTE Cases**<br>**(n=20,359)** | **Comparators**<br>**(n=203,590)** | **Total**<br>**(n=223,949)** |
| --- | --- | --- | --- |
| **Region** |  |  |  |
| 1 Northeast | 8.28% (1686) | 8.85% (18010) | 8.79% (19696) |
| 2 Midwest | 27.81% (5661) | 25.55% (52021) | 25.76% (57682) |
| 3 South | 43.77% (8912) | 44.77% (91138) | 44.68% (100050) |
| 4 West | 19.98% (4067) | 20.11% (40947) | 20.10% (45014) |
| 5 Unknown | 0.16% (33) | 0.72% (1474) | 0.67% (1507) |
| **Age\*** |  |  |  |
| 50-55 | 33.35% (6789) | 34.84% (70927) | 34.70% (77716) |
| 56-60 | 34.80% (7084) | 33.52% (68237) | 33.63% (75321) |
| 61-65 | 31.86% (6486) | 31.64% (64426) | 31.66% (70912) |
| **EH Comorbidities** |  |  |  |
| None | 13.81% (2812) | 37.32% (75981) | 35.18% (78793) |
| 1-2 | 30.25% (6159) | 40.23% (81909) | 39.32% (88068) |
| 3+ | 55.94% (11389) | 22.45% (45700) | 25.49% (57089) |
| **Cancer** |  |  |  |
| None | 74.00% (15065) | 94.42% (192220) | 92.56% (207285) |
| Non-metastatic | 11.11% (2262) | 4.80% (9764) | 5.37%, (12026) |
| Metastatic | 14.89% (3032) | 0.79% (1606) | 2.07%, (4638) |
| **Hospital/Surgery** | 30.22%, (6152) | 2.29%, (4672) | 4.83%, (10824) |
| **Trauma** | 15.03%, (3060) | 2.96%, (6030) | 4.06%, (9090) |
| **Varicose Veins** | 2.73% (556) | 0.88%, (1790) | 1.05%, (2346) |
| **Any hypercoag.** | 10.99%, (2237) | 1.25%, (2551) | 2.14%, (4788) |
| **CAD** | 13.47% (2743) | 4.80% (9775) | 5.59% (12518) |
| **Stroke** | 7.34% (1495) | 2.40% (4886) | 2.85% (6381) |
| **Smoking** | 23.13% (4710) | 9.46% (19263) | 10.70% (23973) |

\*Cases and controls were matched on age ± two years.

### HT Recency of Exposure

Because VTE risk diminished within 60 days of discontinuation, “current” use was defined as hormone exposure within 0-60 days of the index date. Within the previous year, hormone exposures occurred in 11.19% (25,053/223,949) of women; most were continuous users (96.54%) with few initiating therapy within 60 days. Within the most recent 60 days, 8.73% (10.46% cases, 8.56% controls) were exposed. Those with a filled-prescription resulting in exposure within 30 days of the index date had an odds ratio of 1.53 (95%CI: 1.45-1.62) compared to those without hormone exposure in the past year (Table 2). The odds ratio for those without a filled-prescription in the previous 30 days but who had filled a prescription resulting in exposure within 31-60 days was elevated (OR=1.22, 95%CI: 0.99-1.50), but not significantly different from no exposure. The odds ratio for a 0-30 day exposure was 26% higher than for the 31-60 day exposure (OR=1.26, 1.02-1.56) and the odds ratio for the 31-60 day exposure was 35% higher than for 61-90 day exposure (OR=1.35, 0.95-1.92). Odds for exposures 61-90 days prior to the index date were not elevated compared to the past 91-365 days (OR=1.04, 0.77-1.41) or compared to no exposure in the past year (OR=0.90, 0.68,1.19). Thus, current exposure was defined as any 0-60 day exposure and “past” use was defined as any exposure in the previous 61-365 days. The odds ratio for current exposure to *any* estrogen or progestogen was 1.51 (95% CI: 1.42, 1.59) compared to no exposure. Current users tended to be slightly younger (44% of exposed and 34% unexposed were 50-55 years).

**TABLE 2:** ODDS RATIOS FOR VTE BY RECENCY OF EXPOSURE.

| **Last Exposure** | **Adj. OR<sup>a</sup>**<br>**(95% CI)** | **Adj. OR<sup>a</sup>**<br>**(95% CI)** |
| --- | --- | --- |
| **“Current”** | 1.53 (1.45,1.62)<br>(0-30d) | 1.51 (1.43,1.59)<br>(0-60d) |
| **31-60d (no 0-31d)** | 1.22 (0.98,1.48) | -- |
| **61-90d (no 0-60d)** | 0.90 (0.68,1.19) | -- |
| **Past** | 0.94 (0.83,1.06)<br>(91-365d, no 0-90d) | 0.93 (0.83, 1.04)<br>(61-365d, no 0-60d) |

a. Adjusted for all variables in Table 1. Reference category is no exposure in past year.

### Type of HT Exposure

Although VTE risk was elevated with any current hormone exposure, risk varied by type of exposure (Table 3). The odds ratio for current MHT exposure with unopposed estrogen was 1.13 (95% CI: 1.04, 1.23; exposed n=9650) and 1.22 (95% CI: 1.10, 1.38; n=5160) with estrogen-progestogen combinations. Current exposure from estrogen-progestogen contraceptives had the strongest odds ratio (OR=5.22, 95% CI: 4.67, 5.84; n=2476), which was four times higher than combined MHT exposures (OR=4.24, 95% CI: 3.64, 4.98). The odds ratio for current use of progestogens alone was 1.49 (95% CI: 1.24, 1.78; n=1486) and 0.49 (95% CI: 0.34, 0.71; n=786) for estrogen-testosterone (with or without a progestogen).

**TABLE 3:** ODDS RATIOS FOR VTE FROM VARIOUS HORMONE EXPOSURES.

| **HORMONAL EXPOSURES** | **N** | **Controls**<br>**(n=203,590)** | **Cases**<br>**(n=20,359)** | **Adjusted ORs <sup>a</sup>**<br>**(95% CI)** |
| --- | --- | --- | --- | --- |
| **ANY current** estrogen or progestogen | 19558 | 17428 | 2130 | 1.51 (1.43, 1.59) |
| **Estrogen only (MHT)** | 9650 | 8735 | 915 | 1.13 (1.04, 1.23) |
| Oral | 6638 | 5907 | 731 | 1.33 (1.21, 1.46) |
| Estradiol | 3680 | 3294 | 386 | 1.24 (1.09, 1.40) |
| CEE | 2746 | 2426 | 320 | 1.46 (1.28, 1.68) |
| Other estrogens | 212 | 187 | 75 | 1.39 (0.87, 2.24) |
| Transdermal/estradiol | 2885 | 2713 | 172 | 0.70 (0.59, 0.84) |
| Vaginal/estradiol ester<sup>b</sup> | 106 | 97 | 9 | 1.04 (0.44, 2.46) |
| IM/Estrogen<sup>c</sup> | 21 | 18 | 3 | 0.38 (0.08, 1.77) |
| **Estrogen+Progestogens (MHT)** | 5160 | 4727 | 433 | 1.22 (1.10, 1.38) |
| Oral | 3947 | 3582 | 365 | 1.40 (1.24, 1.59) |
| Estradiol+Any Progest | 1995 | 1844 | 151 | 1.14 (0.95, 1.37) |
| Estradiol+Norethindrone | 1014 | 945 | 69 | 1.03 (0.78, 1.35) |
| Estradio+other | 981 | 899 | 82 | 1.25 (0.97, 1.61) |
| CEE/MPA<sup>d</sup> | 1519 | 1363 | 156 | 1.52 (1.25, 1.84) |
| Ethinyl estradiol+norethindrone | 413 | 358 | 55 | 2.35 (1.71, 3.25) |
| Other estrogens/other Progest | 20 | 17 | 3 | 1.33 (0.25, 7.14) |
| Transdermal/estradiol+Progest<sup>e</sup> | 1213 | 1145 | 68 | 0.73 (0.55, 0.96) |
| +Progesterone | 487 | 465 | 22 | 0.55 (0.34, 0.89) |
| +Micronized Progesterone | 262 | 249 | 13 | 0.63 (0.34, 1.19) |
| + Norethindrone | 304 | 288 | 16 | 0.65 (0.36, 1.14) |
| +MPA | 55 | 49 | 6 | 1.76 (0.69, 4.50) |
| + other | 105 | 94 | 11 | 1.74 (0.87, 3.46) |
| **Estrogen+Progest (Contraception)** | 2476 | 1915 | 561 | 5.22 (4.67, 5.84) |
| Oral/ethinyl estradiol<sup>f</sup> | 2399 | 1862 | 537 | 5.11 (4.57, 5.73) |
| Trans/ethinyl estradiol | 13 | 11 | 2 | 5.34 (1.06, 26.79) |
| Vag/ethinyl estradiol | 64 | 42 | 22 | 9.26 (5.31, 16.15) |
| **Estrogen/CEE+Testosterone (±P)<sup>g</sup>** | 786 | 745 | 41 | 0.49 (0.34, 0.71) |
| **Progestogen only** | 1486 | 1306 | 180 | 1.48 (1.24, 1.78) |
| Progesterone | 588 | 550 | 38 | 0.83 (0.57, 1.20) |
| Micronized Progesterone | 223 | 201 | 22 | 1.20 (0.73, 1.97) |
| Norethindrone<sup>h</sup> | 232 | 198 | 34 | 1.99 (1.30, 3.03) |
| MPA | 443 | 357 | 86 | 2.20 (1.67, 2.92) |
| ANY Past Estrogen or Progest (61-365d) | 5494 | 4995 | 499 | 0.94 (0.84, 1.05) |
| NO exposure (ref) | 198,897 | 181,167 | 17,730 | 1.00 |

a. Models adjusted for all risk factors in Table 1 and reference is no hormone use in past year (n=198,897).
b. Vaginal estrogen only (FemRing) also includes those with oral progestogen (n=17).
c. Includes IM estrogen+progestogen (n=1)
d. Includes other progestogens (n=98)
e. Includes transdermal estrogen-progestogen and transdermal estrogen with oral progestogen.
f. Includes oral mestranol (n=6), estradiol+norgestimate with levonorgestrel (plan B) (n=1), and estradiol ester (n=1).
g. Includes some with progestogens (n=146).
h. Includes other progestogens (n=1)

For MHT, oral exposures had higher risk than transdermal or vaginal exposures compared to unexposed. The odds ratio for oral, unopposed estrogen was 1.33 (95% CI: 1.21, 1.46; n=6638) and 1.40 (95% CI: 1.24, 1.59; n=3947) for oral estrogen-progestogen combinations. Transdermal estradiol, whether unopposed (OR=0.70, 95% CI: 0.59, 0.84; n=2885) or with a progestogen (OR=0.73, 95% CI: 0.55, 0.96; n=1213) had the lowest risk. Transdermal estradiol-progestogen (n=380) and transdermal estradiol with an oral progestogen (n=833) were combined as neither was significantly different from no exposure nor from one another. Odds ratios for oral exposures were almost twice as high as transdermal exposures (estrogen only OR=1.90, 95% CI: 1.56, 2.32; combined therapy OR=1.92, 95% CI: 1.43, 2.60). Vaginal exposures (FemRing) also may be low-risk (OR=1.04, 95% CI: 0.44, 2.46; n=106), but the subsample was small. Categories for menopausal vaginal exposures with unopposed estrogen (n=89) and those supplemented with an oral progestogen (n=17) were combined as neither was significantly different from no exposure nor from one another.

Estrogen MHT formulation also affected risk. With oral unopposed estrogen, the odds ratio was higher with CEE (OR=1.46, 95% CI: 1.28, 1.68; n=2746) than estradiol (OR=1.24, 95% CI: 1.09, 1.40; n=3,680), although not significantly so (OR=1.19, 95% CI: 0.99, 1.43). Among women exposed to oral estrogen, for each 2320 women taking estradiol or 1211 taking CEE, one additional woman will experience a VTE. Odds ratios for oral estrogen-progestogen combinations were lowest with estradiol (OR=1.14, 95% CI: 0.95, 1.37; n=1995), higher with CEE (OR=1.52, 95% CI: 1.25, 1.84; n=1519), and highest with ethinyl estradiol (OR=2.35, 95% CI: 1.71, 3.25; n=413) compared to no hormone exposure. The odds ratio was significantly higher for CEE compared to estradiol (OR=1.33, 95% CI: 1.02, 1.72) and for ethinyl estradiol compared to CEE (OR=1.55, 95% CI: 1.07, 2.25). Ethinyl estradiol-norethindrone more than doubled the odds of a VTE, but estradiol-norethindrone was not associated with VTE. Among women exposed to oral combined MHT, for each 3,976 women taking estradiol+progestogen (NNH=3976), 1071 women taking CEE+progestin (NNH=1071), or 413 women taking ethinyl estradiol+progestin (NNH=413), one additional woman will experience a VTE.

Risk was very high for contraceptive exposures (almost exclusively ethinyl estradiol+progestin combinations) compared to no hormone exposure. Contraceptive users tended to be younger (94% were 50-54 years) and most had oral exposures (OR=5.11, 95% CI: 4.57, 5.73; n=2399). Transdermal exposures did not have lower risk (OR=5.34, 95% CI: 1.06, 26.78; n=13) and risk for vaginal contraceptives was even higher (NuvaRing, OR=9.26, 95% CI: 5.31, 16.15; n=64). For each 132 women exposed to estrogen-progestogen contraceptives, one additional woman will experience a VTE (NNH=132).

Because there were many progestogens with small subgroup sizes, we present – for descriptive purposes only - the three categories most sensitive to physician choice: progestogens combined with oral estradiol or with transdermal estradiol, and progestogens alone. Oral estradiol was most often combined with norethindrone (n=1014), but also with: MPA (n=465), progesterone (n=280), micronized progesterone (n=143), levonorgestrel (n=1), norgestimate (n=38), and drospirenone (n=56). Oral CEE was most often combined with MPA (n=1,519).

When progestogens were given alone, odds were highest for MPA (OR=2.20, 95% CI: 1.67, 2.92; n=443), followed by norethindrone (OR=1.99, 95% CI: 1.30, 3.03; n=232), micronized progesterone (OR=1.20, 95% CI: 0.73, 1.97; n=223), and progesterone (OR=0.83, 95% CI: 0.57, 1.20; n=588). A similar ordering was observed when progestogens were combined with transdermal estradiol: VTE odds were highest with MPA and lowest with progesterone.

Sub-analyses checked model stability and robustness across subgroups. When those with cancer or a hypercoagulable condition (n=15,156 cancer only, n=3280 hypercoagulability only, n=1508 both) were excluded, results from the full sample were similar for MHT exposures, but showed some increases for CEE and contraceptive exposures (Table 4). When stratified on age (<58 and ≥58), transdermal exposures remained without elevated risk across subsamples, but VTE risk from MHT was slightly higher in younger women. In contrast, risk from contraceptive exposures was markedly elevated in older women (OR=8.95, 95% CI: 4.59, 17.44) than in younger women (OR=4.83, 95% CI: 4.31, 5.41).

**TABLE 4:** SUBSAMPLE RESULTS FOCUSING ON ROUTE AND FORMULATION OF ESTROGEN EXPOSURES.

| **HORMONAL EXPOSURES** | **TOTAL SAMPLE<sup>a</sup>** | **SUBSAMPLE:**<br>**NO CANCER, NO**<br>**HYPERCOAG.<sup>b</sup>** | **SUBSAMPLE:**<br>**AGE<58<sup>a</sup>** | **SUBSAMPLE:**<br>**Age ≥58<sup>a</sup>** |
| --- | --- | --- | --- | --- |
| **Estrogen only (MHT)** |  |  |  |  |
| Oral |  |  |  |  |
| Estradiol | 1.24 (1.09, 1.40) | 1.27 (1.11, 1.46) | 1.54 (1.29, 1.84) | 1.17 (0.98, 1.39) |
| CEE | 1.46 (1.28, 1.68) | 1.53 (1.32, 1.78) | 1.75 (1.42, 2.16) | 1.41 (1.17, 1.69) |
| Other estrogens | 1.39 (0.87, 2.24) | 1.52 (0.91, 2.53) | 1.08 (0.42, 2.77) | 1.87 (1.08, 3.25) |
| Transdermal/estradiol | 0.70 (0.59, 0.83) | 0.69 (0.57, 0.84) | 0.78 (0.61, 0.99) | 0.63 (0.48, 0.81) |
| Vaginal/estradiol ester<sup>c</sup> | 0.96 (0.43, 2.14) | 0.87 (0.36, 2.07) | 1.46 (0.48, 4.44) | 0.62 (0.18, 2.13) |
| IM/Estrogen<sup>d</sup> | 0.38 (0.08, 1.77) | 0.34 (0.06, 2.11) | 0.79 (0.07, 8.46) | 0.31 (0.04, 2.23) |
| **Estrogen+Progest (MHT)** |  |  |  |  |
| Oral |  |  |  |  |
| Estradiol | 1.14 (0.95, 1.37) | 1.18 (0.97, 1.45) | 1.14 (0.88, 1.47) | 0.97 (0.74, 1.28) |
| CEE/MPA<sup>e</sup> | 1.52 (1.25, 1.84) | 1.69 (1.38, 2.07) | 1.55 (1.18, 2.04) | 1.38 (1.06, 1.80) |
| Ethinyl estradiol+norethindrone | 2.35 (1.71, 3.25) | 2.41 (1.70, 3.41) | 2.13 (1.33, 3.43) | 2.11 (1.36, 3.30) |
| Other estrogens | 1.33 (0.25, 7.14) | 1.67 (0.27, 10.36) | 1.40 (0.06, 35.21) | 1.07 (0.15, 7.59) |
| Transdermal/estradiol<sup>f</sup> | 0.73 (0.56, 0.96) | 0.70 (0.51, 0.96) | 0.62 (0.42, 0.92) | 0.78 (0.53, 1.15) |
| **Estrogen+Progest (contraceptives)** | 5.22 (4.67, 5.83) | 5.59 (4.96, 6.30) | 4.83 (4.31, 5.41) | 8.95 (4.59, 17.44) |
| **Estrogen/CEE+Testosterone (±P)<sup>g</sup>** | 0.49 (0.34, 0.71) | 0.46 (0.30, 0.71) | 0.49 (0.28, 0.85) | 0.46 (0.28, 0.77) |
| **Progestogens only** | 1.48 (1.23, 1.78) | 1.45 (1.18, 1.77) | 1.70 (1.37, 2.12) | 1.29 (0.92, 1.81) |
| Past HT (61-365d) | 0.94 (0.84, 1.05) | 0.88 (0.77, 1.01) | 1.01 (0.87, 1.17) | 0.95 (0.80, 1.14) |

a. Models adjusted for all risk factors in Table 1 and reference is no use in past year.
b. Models adjusted for risk factors in Table 1, except cancer and hypercoaguable conditions; reference is no use in past year.
c. Vaginal estrogen only (FemRing) also includes some with oral progestogen;
d. May include IM estrogen+progestogen (n=1)
e. May include other progestogens (n=98)
f. May include transdermal EP and transdermal E with oral P;
g. May include progestogens (n=146)

## DISCUSSION

### Principal findings

Since the landmark trials revealed the potential for thrombotic risk with MHT, large observational studies have provided convincing evidence that transdermal estradiol, whether unopposed or combined with a progestogen, did not increase VTE risk, and for oral therapy, estradiol was safer than CEE. This study strengthens that evidence and uniquely adds information regarding risk within the context of US prescribing patterns. In this study, results indicated that more than two-thirds of transdermal estradiol exposures were combined with an oral progestogen. Transdermal exposures remained lower risk, but risk may increase when combined with a higher risk oral progestin (e.g., MPA). In addition, MHT with ethinyl estradiol may be more common in the US than in Europe and increased risk more than CEE. VTE risk, however, appeared to diminish within 60 days of hormone discontinuation.

A striking finding in this study was the five-fold increase in VTE risk with estrogen-progestin contraceptives compared to no hormone exposure (NNH=132) and a four-fold increase compared to MHTs. In other studies and depending upon the progestin, oral combined contraceptives have had an approximate four-fold increase in VTE risk for women 15-50 years of age[27,28] and a six-fold increase in women 50 and older.[11] In this study, there was almost a nine-fold increase in women over 58. Whether contraceptives increased risk because of dosage, hormone formulation, or older age is unclear.

### Strengths and weaknesses

A principal limitation of this study is that insurance claims data may not capture important information. Some risk factors (e.g., smoking) are challenging to accurately capture in claims data and may be underestimated. We lacked detailed personal information, such as socioeconomic status and age at menopause. Exposure was estimated from filled prescriptions without information on adherence, although most women who filled hormone prescriptions did so continuously. We also lacked data on the indication for prescriptions, particularly limiting interpretation for women on progestogen-only regimens, as potential indications range from contraception to abnormal uterine bleeding to menopausal symptoms. While these limitations may result in some residual confounding, it is unlikely that they would be systematically different between cases and controls.

Results of this study may be limited by the observational design. Randomization in clinical trials alleviates bias and confounding, but such trials are costly, time consuming, and unlikely to be large enough to test various formulations and routes of exposures. Insurance claims data provide sufficiently large samples for more nuanced analyses. Also, the nested case-control design used in this study with a time-restricted approach offers a strong and efficient method that results in unbiased estimation of exposure risk for the population in the base cohort. We statistically controlled for factors associated with the outcome variable (VTE risk factors) and factors likely to be associated with exposure (CAD, stroke, and the Elixhauser index) to minimize indication bias. Nevertheless, results, especially possible protective effects from transdermal estradiol or estrogen+testosterone, are suggestive until tested in a clinical trial.

Primary data collection efforts assessing thrombotic risk due to MHT have had limited sample sizes to explore exposure routes and formulations. VTE incidence is infrequent (129/100,000)[18] and large medical record studies and meta-analyses have facilitated assessment of risk from various types of exposures. VTE risks in this study are consistent with those from a recent meta-analysis[26] and large UK clinical databases[13] for oral unopposed estrogen and transdermal estrogen. For oral combined MHT, the meta-analysis found heterogeneity across studies. Our results were similar to UK results[13], but we used a more restrictive VTE definition and age range and found lower risk for oral CEE+MPA. Our results also suggested that transdermal VTE risk may increase when combined with higher risk oral progestins, such as MPA. Progestogens and specifically MPA can increase the thrombotic risk of estrogens.[10–13] In this study, MPA had the highest risk, while the effect of oral norethindrone was unclear. In the UK study, progestogen-only exposures had higher risk than in this study, possibly due to differences in prescribed progestogens.

### Implications and future research

This is the first large, detailed study on hormone risk in US women 50-64 years of age. The study confirms prior findings that estradiol, and in particular transdermal estradiol, offers MHT with lower risk for VTE. However, more research is needed on the effects of various oral progestogens when combined with transdermal estradiol, as two-thirds of the transdermal estradiol exposures were combined with oral progestogens. In addition, results suggest that ethinyl estradiol carries an even higher VTE risk than CEE. A striking finding was the four-fold higher VTE risk from contraceptive exposures compared to MHT for women 50 years of age and older. The markedly increased VTE risk from combined hormonal contraceptives – whether oral, transdermal, or vaginal – suggests that they should not be used for peri-menopausal symptoms without need for the contraceptive effect. Women should be counselled on the risks of continuing hormonal contraceptives into older ages and using hormonal contraceptives for MHT. Further research needs to explore hormonal risks across a wider age range from the same cohort, including women over 50 for contraceptive exposures and women under 50 for MHT exposures.

## Supporting information

STROBE checklist

## Data Availability

Data availability statement
Data were obtained from a third party and are not publicly available. All data relevant to the study are summarised in the article. No further data are available. Data are proprietary of Optum Health Systems. However, requests for re-analyses will be considered.

## ACKNOWLEDGEMENTS

The authors would like gratefully to acknowledge Joel Weissfeld, MD, MPH, at the FDA for sharing his protocol and hormone list. We also thank the Texas Academy of Family Physicians for funding.

